# Identification of potential biomarkers and inhibitors for SARS-CoV-2 infection

**DOI:** 10.1101/2020.09.15.20195487

**Authors:** Hanming Gu, Gongsheng Yuan

## Abstract

The COVID-19 pandemic caused by severe acute respiratory syndrome coronavirus 2 (SARS-CoV-2) has overwhelmed many health systems globally. Here, we aim to identify biological markers and associated biological processes of COVID-19 using a bioinformatics approach to elucidate their potential pathogenesis. The gene expression profile of the GSE152418 dataset was originally produced by using the high-throughput Illumina NovaSeq 6000. Kyoto Encyclopedia of Genes and Genomes pathway (KEGG) and Gene Ontology (GO) enrichment analyses were applied to identify functional categories and biochemical pathways. KEGG and GO results suggested that biological pathways such as “Cancer pathways” and “Insulin pathways” were mostly affected in the development of COVID-19. Moreover, we identified several genes including EP300, CREBBP, and POLR2A were involved in the virus activities in COVID-19 patients. We further predicted that some inhibitors may have the potential to block the SARS-CoV-2 infection based on the L1000FWD analysis. Therefore, our study provides further insights into the underlying pathogenesis of COVID-19.

## Introduction

COVID-19 caused by SARS-CoV-2 infection, has affected a large number of countries with increasing morbidity and mortality^1^. Most COVID-19 patients exhibited mild-to-moderate symptoms and small groups of patients typically within a week turned into a severe stage. Early reports showed 21% of COVID-19 patients died in New York City during March 2020^2^. The aged patients or those with medical comorbidities such as diabetes, hypertension, lung diseases and cardiovascular diseases have a higher mortality rate^3^. Currently, there are no curative therapies for COVID-19. Therefore, understandings of the SARS-CoV-2 pathogenesis are critical to the development of therapeutics.

Recent studies have suggested that uncontrolled inflammation leads to disease severity in COVID-19^4^. Most COVID-19 patients are characterized by increased numbers of neutrophils and exhibit increased levels of pro-inflammatory cytokines including IL6, IL1 and MCP-1 in the plasma^5^. The uncontrolled pro-inflammatory cytokines may lead to shock, respiratory failure and multiple organ failure in COVID-19 patients^6^. However, little is known about the mechanisms underlying COVID-19, and whether individuals in different parts of the world respond differently to SARS-CoV-2 remains unknown.

The SARS-CoV-2 is an RNA virus with spike-like glycoproteins^7^. The development of vaccines for COVID-19 patients are largely dependent on the specific RNA and protein structure^8, 9^. Modern antiviral drug discovery is expected to be impacted dramatically by analyzing genomics^10^. High-throughput microarray methodologies and advanced drug development such as remdesivir have drawn extensive attentions^11^. Thus, there is an urgent need to identify potential targets or biomarkers for COVID-19 patients by genomics.

In this study, we investigated the effect of SARS-CoV-2 on human peripheral blood mononuclear cells (PBMCs). We analyzed and identified several DEGs, inhibitors and the relevant biological processes of COVID-19 by utilizing comprehensive bioinformatics analysis. We performed the functional enrichment, pathway analysis, and protein-protein interaction for finding COVID-19 gene nodes in PBMCs. These key genes and pathogenetic factors could be critical to guide future clinical and therapeutic interventions.

## Methods

### Data resources

Gene expression profile dataset GSE152418 was obtained from the GEO database (http://www.ncbi.nlm.nih.gov/geo/). The data was produced by using an Illumina NovaSeq 6000 (Homo sapiens) (Developmental and Cognitive Neuroscience, Yerkes National Primate Research Center, Atlanta, GA30329-4208, US). The GSE152418 dataset contained data including 17 COVID-19 subjects and 17 healthy controls.

### Data acquisition and preprocessing

The raw microarray data between SARS-CoV-2 positive samples and negative controls were subsequently conducted by R script. We used a classical t test to identify DEGs with P<.01 and fold change ≥1.5 as being statistically significant.

### Gene Ontology (GO) and pathway enrichment analysis

Gene Ontology (GO) analysis is a widely used approach to annotate genomic data and identify characteristic biological information. The Kyoto Encyclopedia of Genes and Genomes (KEGG) database is commonly used for systematic analysis of gene functions and annotation of biological pathways. GO analysis and KEGG pathway enrichment analysis of DEGs in this study were analyzed by the Database for Annotation, Visualization, and Integrated Discovery (DAVID) (http://david.ncifcrf.gov/). P<.05 and gene counts >10 were considered statistically significant.

### Module analysis

The Molecular Complex Detection (MCODE) was used to analyze the densely connected regions in PPI networks. The significant modules were from constructed PPI networks using MCODE. The function and pathway enrichment analyses were performed by using DAVID, and P<.05 was used as the cutoff criterion.

### Inhibitor analysis prediction

The L1000FWD, a NIH Common Fund program, is used to identify potential novel inhibitors. L1000FWD calculated the similarity between an input gene expression signature and the LINCS-L1000 data to rank inhibitors which can regulate the transcriptional signature^12^. The adjusted p-value of 0.05 has been considered as threshold for statistical significance.

### Cell culture and treatment

The RAW 264.7 cell lines were purchased from American Type Culture Collection (ATCC® TIB-71™). Cells were cultured in DMEM medium supplemented with 10% FBS and 1% penicillin/streptomycin and incubated at 37 °C under 5% CO2. The cells were induced with 1µg/mL LPS for 24 hours and treated with different potential inhibitors for 24 hours: anisomycin (20µM, Sigma), BRD-K60870698 (20µM, Santa Cruz), QL-×-138 (20µM, Probechem), BMS-345541 (20µM, Sigma), homoharringtonine (20µM, Sigma) and kinetin-riboside (20µM, Sigma).

### Real-time PCR

The total RNA from cells was extracted using TRIzol reagent (Invitrogen, USA) as described previously^13^. The cDNA was obtained using a reverse transcription kit according to the manufacturer’s instructions (TAKARA, US). PCR amplification was carried out for a total of 40 cycles and normalized to GAPDH expression. All reactions were performed in triplicate, and the relative expression was determined using the 2-ΔΔCT method. The primers used are listed as follows: GAPDH(F):AGGTCGGTGTGAACGGATTTG,GAPDH(R):TGTAGACCATGTAGTTGAGGTCA;IL1(F):GCCCATCCTCTGTGACTCAT,IL1(R):AGGCCACAGGTATTTTGTCG;IL6 (F):AGTTGCCTTCTTGGGACTGA,IL6(R):TCCACGATTTCCCAGAGAAC;TNFα(F):CGTCAGCCGATTTGCTATCT, TNFα(R):CGGACTCCGCAAAGTCTAAG.

### Statistical analysis

For statistical analysis, Prism 7 software (GraphPad Software, USA) was used. The data were expressed as the mean ± S.E.M. A two-tailed Student’s t-test was performed to determine the significance of the difference between the two groups. One-way analysis of variance (ANOVA) with Dunnett’s post hoc test was used to compare more than two groups. P values < 0.05 were considered significant.

## Results

### Identification of DEGs in COVID-19 patients

To gain the insights on host responses to SARS-CoV-2 infection, the modular transcriptional signature of COVID-19 patients was compared to that of the healthy controls. A total of 1254 genes were identified to be differentially expressed in COVID-19 samples with the threshold of P<0.001. The top 10 up- and down-regulated genes for COVID-19 and negative samples are list in Table 1.

**Table 1.**

| **Entrez gene** | **Gene Symble** | **Fold-change** | **Regulation** |
| --- | --- | --- | --- |
| Top 10 down-regulated DEGs in COVID-19 |  |  |  |
| 3006 | H1-2 | -1.524994 | Down |
| 55355 | HJURP | -1.511164 | Down |
| 9700 | ESPL1 | -1.475812 | Down |
| 995 | CDC25C | -1.471294 | Down |
| 24137 | KIF4A | -1.463716 | Down |
| 4288 | MKI67 | -1.459898 | Down |
| 9212 | AURKB | -1.456353 | Down |
| 991 | CDC20 | -1.453053 | Down |
| 80119 | PIF1 | -1.451448 | Down |
| 28468 | IGHV1-18 | -1.451174 | Down |
| Top 10 up-regulated DEGs in COVID-19 |  |  |  |
| 10016 | PDCD6 | 1.802808 | up |
| 54778 | RNF111 | 1.763973 | up |
| 152006 | RNF38 | 1.762005 | up |
| 9807 | IP6K1 | 1.748826 | up |
| 81566 | CSRNP2 | 1.744855 | up |
| 7048 | TGFBR2 | 1.734719 | up |
| 91526 | ANKRD44 | 1.729817 | up |
| 9844 | ELMO1 | 1.719519 | up |
| 85364 | ZCCHC3 | 1.719157 | up |
| 55810 | FOXJ2 | 1.718367 | up |

### Enrichment analysis of DEGs in COVID-19 patients

To further analyze the biological roles and potential mechanisms of DEGs from the COVID-19 samples versus healthy controls, we performed KEGG pathway and GO categories enrichment analysis. KEGG pathways (http://www.genome.jp/kegg/) is an encyclopedia of genes and genomes for understanding high-level functions. Our study showed top ten enriched KEGG pathways including “Pathways in cancer”, “Insulin signaling pathway”, “Neurotrophin signaling pathway”, “T cell receptor signaling pathway”, “Fc gamma R-mediated phagocytosis”, “Pancreatic cancer”, “Phosphatidylinositol signaling system”, “Inositol phosphate metabolism”, “Acute myeloid leukemia”, and “Chronic myeloid leukemia” (Figure 1).

**Figure 1.**
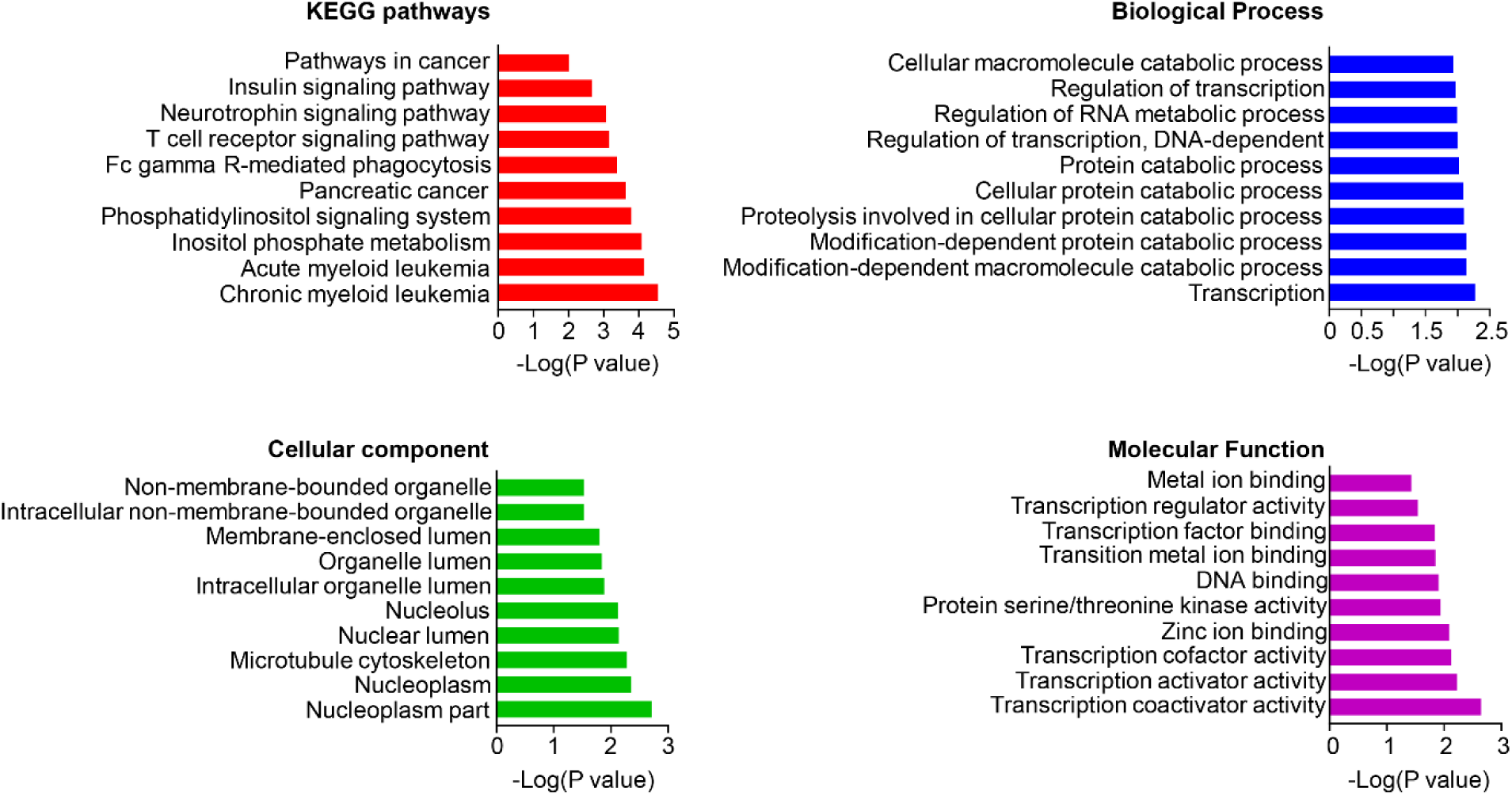
The KEGG pathways, biological process, cellular component, and molecular function terms enriched by the DEGs between COVID-19 patients and healthy controls. DEGs =differentially expressed genes, KEGG = Kyoto Encyclopedia of Genes and Genomes.

Gene Ontology (GO) analysis is a major bioinformatic initiative to unify the representation of gene and gene product, which includes cellular components, molecular functions, and biological processes. We identified top ten cellular components including “Non-membrane-bounded organelle”, “Intracellular non-membrane-bounded organelle”, “Membrane-enclosed lumen”, “Organelle lumen”, “Intracellular organelle lumen”, “Nucleolus”, “Nuclear lumen”, “microtubule cytoskeleton”, “Nucleoplasm”, and “Nucleoplasm part” (Figure 1). We then identified top ten biological processes: “Cellular macromolecule catabolic process”, “Regulation of transcription”, “Regulation of RNA metabolic process”, “Regulation of transcription, DNA-dependent”, “Protein catabolic”, “Cellular protein catabolic process”, “Proteolysis involved in cellular protein catabolic process”, “Modification-dependent protein catabolic process”, “Modification-dependent macromolecule catabolic process”, and “Transcription” (Figure 1). We also identified top ten molecular functions: “Metal ion binding”, “Transcription regulator activity”, “Transcription factor binding”, “Transition metal ion binding”, “DNA binding”, “Protein serine/threonine kinase activity”, “Zinc ion binding”, “Transcription cofactor activity”, “Transcription activator activity”, and “Transcription coactivator activity” (Figure 1).

### PPI network and Module analysis

The PPI network was constructed to further explore the relationships of DGEs at the protein level. We set the criterion of combined score >0.7 and created the PPI network by using the 1198 nodes and 3137 interactions between negative controls and COVID-19 positive samples. Among these nodes, the top ten genes with highest scores are shown in Table 2.

**Table 2.** Top ten genes demonstrated by connectivity degree in the PPI network.

| Gene Symbol | Gene title | Degree |
| --- | --- | --- |
| EP300 | E1A binding protein p300 | 70 |
| CDC20 | Cell division cycle 20 | 61 |
| UBE2C | Ubiquitin conjugating enzyme E2 C | 61 |
| CREBBP | CREB binding protein | 56 |
| POLR2A | RNA polymerase II subunit A | 55 |
| BTRC | Beta-transducin repeat containing E3 ubiquitin protein ligase | 55 |
| ATG7 | Autophagy related 7 | 54 |
| CUL3 | Cullin 3 | 54 |
| FBXW11 | F-box and WD repeat domain containing 11 | 50 |
| DCAF1 | DDB1 and CUL4 associated factor 1 | 48 |

The top two significant modules of COVID-19 versus control samples were selected to depict the functional annotation of genes (Figure 2). We identified top ten signaling pathways in module 1: Antigen processing, Neddylation, Class I MHC mediated antigen processing and presentation, Adaptive Immune System, Immune System, Post-translational protein modification, Metabolism of proteins, MAP3K8 (TPL2)-dependent MAPK1/3 activation, Conversion from APC/C:Cdc20 to APC/C:Cdh1 and Inhibition of the proteolytic activity of APC/C. We also identified top ten signaling pathways in module 2: mRNA Splicing (Major), mRNA Splicing, RNA polymerase II transcribes snRNA genes, Processing of Capped Intron-Containing Pre-mRNA, Metabolism of RNA, mRNA Splicing (Minor), RNA Polymerase II Transcription, Transcription of the HIV genome, Gene expression (Transcription), RNA Polymerase II Pre-transcription Events by using Reactome Pathway Database (https://reactome.org/) (Supplemental Table S1).

**Figure 2.**
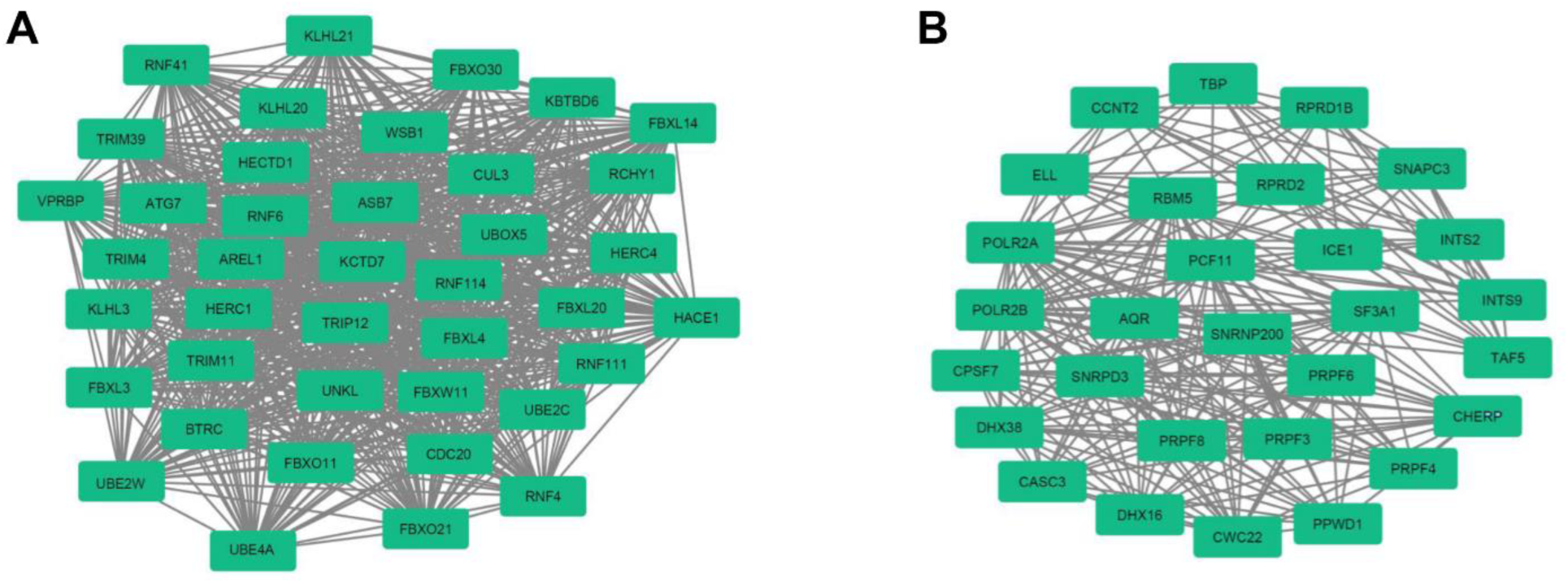
Top 2 modules from the protein-protein interaction network between COVID-19 patients and healthy controls.

### Potential inhibitors for COVID-19 disease

We highlighted top ten inhibitors (Table 3) and further selected six potential anti-COVID-19 inhibitors with the highest scores identified by using the L1000FWD analysis (Figure 3). Among them: anisomycin is used as a DNA synthesis inhibitor; BRD-K60870698 is a protein synthesis inhibitor; QL-×-138 is used for PARP inhibition; BMS-345541 is used as an IKK inhibitor, homoharrringtonine is used as a protein synthesis inhibitor; kinetin-riboside is used for EGFR inhibition. Macrophages are key players during SARS-CoV-2 infection in innate immunity: they produce cytokines and lead to the activation and regulation of immune response^14, 15^. We then determined the anti-inflammatory effects of the six predicted inhibitors by using the LPS induced macrophages (Figure 4). We found the anisomycin, QL-×-138 and BMS-345541 can inhibit the IL1, IL6 and TNFα expressions during the LPS induction (Figure 4). It is suggested that the potential inhibitors, anisomycin, QL-×-138, and BMS-345541, may block the SARS-CoV-2 infection and inflammation in COVID-19 patients.

**Table 3.** Potential inhibitors identified by the L1000FWD analysis.

| Inhibitors | Similarity score | p-value | Combined score | Targets |
| --- | --- | --- | --- | --- |
| anisomycin | 0.0363 | 2.65E-04 | 6.28 | DNA synthesis inhibitor |
| BRD-K60870698 | 0.0363 | 3.40E-04 | 5.74 | Protein synthesis inhibitor |
| QL-X-138 | 0.0375 | 5.40E-04 | 5.58 | PARP inhibitor |
| BMS-345541 | 0.0352 | 8.16E-04 | 5.56 | IKK inhibitor |
| homoharringtonine | 0.034 | 6.30E-04 | 5.56 | Protein synthesis inhibitor |
| kinetin-riboside | 0.0352 | 8.98E-04 | 5.09 | EGFR inhibitor |

**Figure 3.**
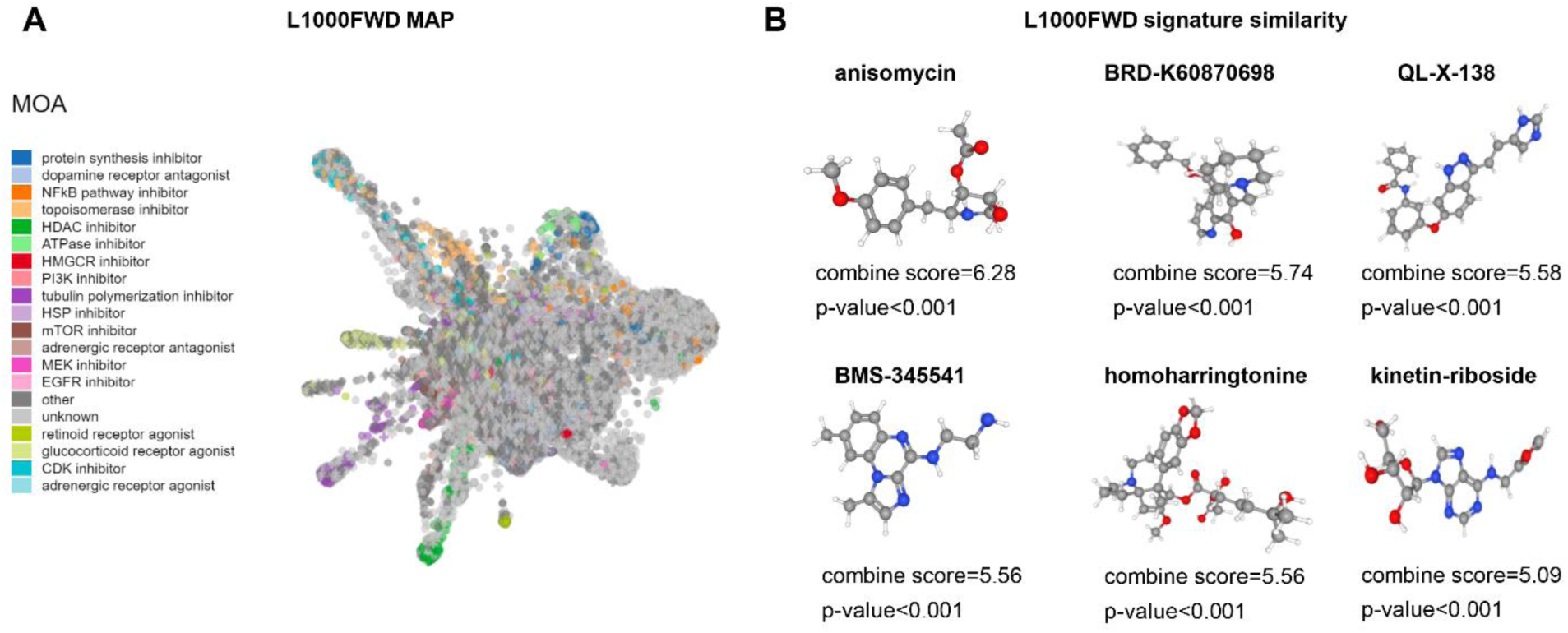
Inhibitors prediction against SARS-CoV-2 infection by L1000FDW visualization. (A) Input genes are represented by the significantly upregulated and downregulated genes obtained from the analysis of the GSE152418 dataset. Dots are color-coded based on the Mode of Action (MOA) of the respective inhibitor. (B) The inhibitors with a high significance p-value and combined score were selected.

**Figure 4.**
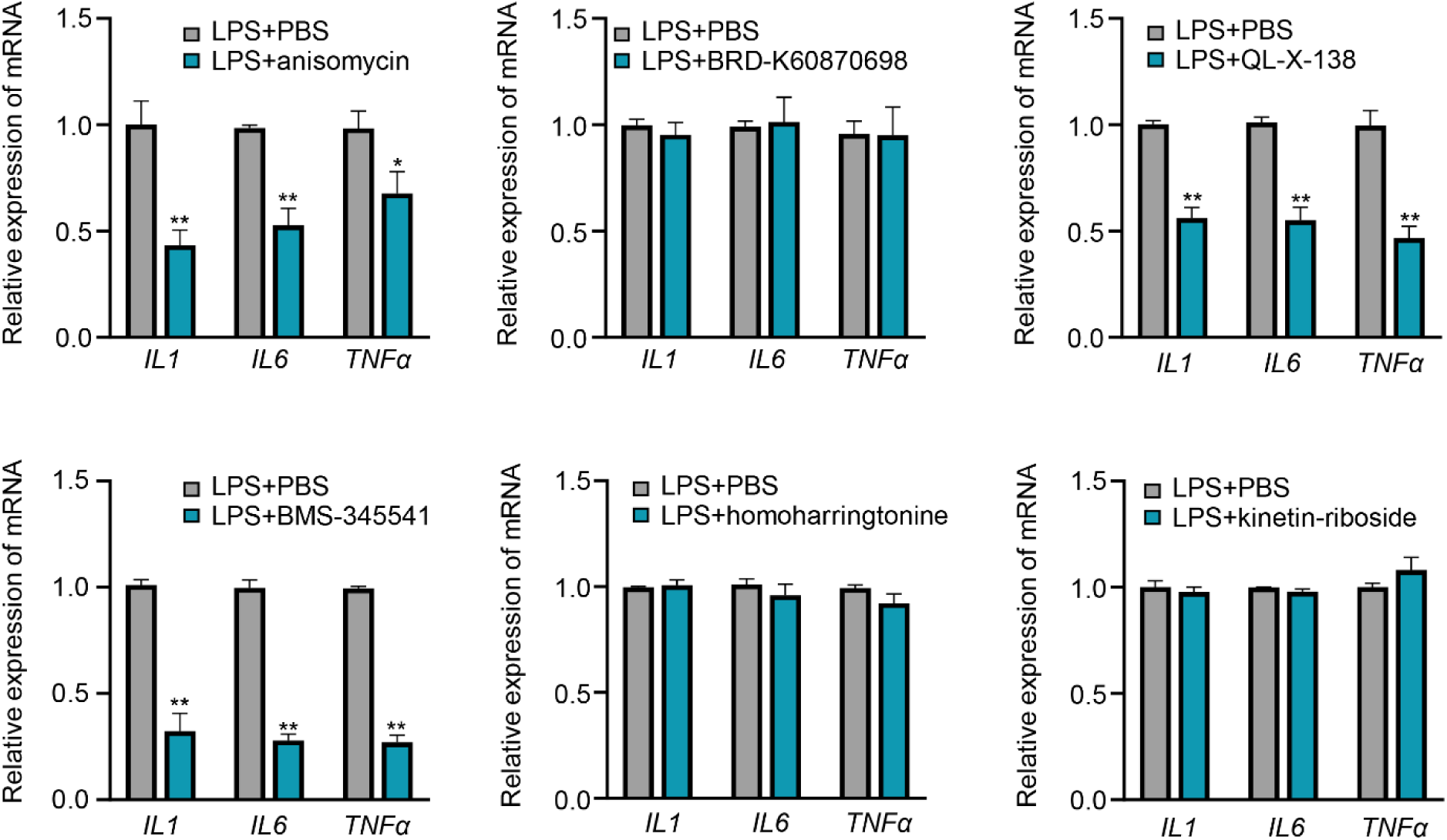
Potential inhibitors decrease the inflammation in macrophages. The RAW264.7 cells were induced with LPS (1µg/mL) for 24 hours and treated with the predicted inhibitors (anisomycin 20µM, BRD-K60870698 20µM, QL-×-138 20µM, BMS-345541 20µM, homoharringtonine 20µM and kinetin-riboside 20µM) for 24 hours. The relative mRNA expression level of pro-inflammatory cytokines IL1, IL6 and TNFα were determined by real-time PCR. **P < 0.01 and *P < 0.05 versus control (n = 5). Note that the anisomycin, QL-×-138 and BMS-345541 depicted anti-inflammation effect in LPS induced macrophages.

## Discussion

The COVID-19 disease caused by the SARS-CoV-2 has turned into a worldwide catastrophe. Understanding the pathogenesis of COVID-19 is highly urgent and critical for diagnosis and treatment.

In the study by Lee LY et al^16^, the phenotype of COVID-19 disease in over half of the cancer patients is mild, but the mortality is higher than that in the general non-cancer population. And based on our KEGG studies, we found the COVID-19 patients were highly relevant to the cancer related pathways including acute myeloid leukemia and chronic myeloid leukemia. Besides cancer, diabetes is also associated with decreased host defense immunity and disordered glucose metabolism, which increases the susceptibility to COVID-19 infection^17^. Additionally, our KEGG studies indicated that COVID-19 took part in the regulation of insulin pathways. It is probable that patients with COVID-19 may aggravate the disorders of insulin and glucose metabolism. Thus, protecting patients with cancer related diseases or diabetes from exposure to SARS-CoV-2 is crucial. Wearing mask, self-isolation, keeping safe distance and avoiding crowded work environments are the best ways to minimize the risk of COVID-19.

In addition, the infection of COVID-19 is also associated with the cellular macromolecule catabolic process including the regulation of RNA metabolic process and the proteolysis involved in cellular protein catabolic process based on our GO analysis. Entry of the SARS-CoV-2 is mainly dependent on proteolytic activation of the spike protein^18^. During the process of viral infection, the spike protein is cleaved into the S1 and S2 subunits and the S2 subunit is released^19^. The other evidence indicated that SARS-CoV-2 S protein can activate protease-independent and receptor-dependent cellular fusion to promote viral spreading^20^.

In our study, the subsequent construction of the PPI network identified several DEGs as potential critical genes during COVID-19 which could be considered as active targets. EP300 and CREBBP target a significant number of proteins for acetylation, including cytosolic proteins involved in essential metabolic processes^21^. POLR2A is a key virus polymerase-interacting protein and is required for viral replication and transcriptional activity^22^. Cell division cycle 20 (CDC20) encodes a regulatory protein and plays important roles in tumorigenesis^23^. Autophagy and the ubiquitin–proteasome system (UPS) are two major intracellular quality control pathways that are responsible for cellular homeostasis in eukaryotes^24^. Here, we identified the UPS related gene UBE2C, BTRC, CUL3, FBXW11, DCAF and autophagy related gene ATG7 were related to SARS-CoV-2 infection.

In our study, we identified a number of anti-COVID-19 inhibitors by using L1000FWD. Interestingly, among these inhibitors, we found that anisomycin, QL-×-138, and BMS-345541 can also block the inflammation in macrophages, which may further inhibit the cytokine storm in COVID-19 disease. Anisomycin inhibits protein synthesis and substantially depresses the levels of the conventional early mRNAs^25^. In the study by V M Quintana et al, anisomycin strongly inhibits the replication of reference strains and clinical isolates of all DENV serotypes and ZIKV virus in Vero cells^26^. QL-×-138 was identified as a selective and potent BTK/MNK dual kinase inhibitor, which exhibited covalent binding to BTK and noncovalent binding to MNK^27^. Nuclear factor-κB (NF-κB) is a critical molecular which is involved in numerous inflammatory processes and diseases^28-30^. The inhibition of NF-κB by BMS-345541 prevents the TNFα-induced rise in PTPN2 protein, independent of apoptotic events^31^. Moreover, BMS-345541 can inhibit the IKKbeta kinase activity from HTLV-1 infected cells^32^.

In summary, we provided the basis for the identification of potential biomarkers for the SARS-CoV-2 infection. Cancer and diabetes are the two major kinds of diseases triggered by SARS-CoV-2 infection. Future studies will focus on single and combined administration of potential anti-SARS-CoV-2 inhibitors in vivo and in vitro studies. Our study thus provides further insights into the mechanism of COVID-19, which may facilitate the diagnosis and treatment.

## Data Availability

Gene expression profile dataset GSE152418 was obtained from the GEO database. http://www.ncbi.nlm.nih.gov/geo/

